# Blood neurofilament light concentration at admittance: a potential prognostic marker in COVID-19

**DOI:** 10.1101/2020.09.07.20189415

**Authors:** Anne Hege Aamodt, Einar August Høgestøl, Trine Haug Popperud, Jan Cato Holter, Anne Ma Dyrhol-Riise, Kristian Tonby, Birgitte Stiksrud, Else Quist-Paulsen, Tone Berge, Andreas Barratt-Due, Pål Aukrust, Lars Heggelund, Kaj Blennow, Henrik Zetterberg, Hanne Flinstad Harbo

**Affiliations:** Oslo University Hospital, Department of Neurology, Oslo, Norway; University of Oslo, Institute of Clinical Medicine, Oslo, Norway; Oslo University Hospital, Department of Microbiology, Oslo, Norway; Oslo University Hospital Department of Infectious Diseases, Oslo University Hospital, Oslo, Norway.; Oslo Metropolitan University, Department of Mechanical, Electronic and Chemical Engineering, Oslo, Norway»; Oslo University Hospital, Department of Research, Innovation and Education, Oslo, Norway; Oslo University Hospital, Division of Emergencies and Critical Care, Rikshospitalet, Oslo, Norway; Oslo University Hospital, Department of Immunology, Oslo University Hospital, Oslo, Norway; Oslo University Hospital, Research Institute of Internal Medicine, Oslo, Norway; Drammen Hospital, Department of Internal Medicine, Vestre Viken Hospital Trust, Drammen, Norway; University of Bergen, Department of Clinical Science, Bergen, Norway; Department of Psychiatry and Neurochemistry, Institute of Neuroscience & Physiology, the Sahlgrenska Academy at the University of Gothenburg, Mölndal, Sweden; Clinical Neurochemistry Laboratory, Sahlgrenska University Hospital, Mölndal, Sweden; Department of Neurodegenerative Disease, UCL Institute of Neurology, London, United Kingdom; UK Dementia Research Institute at UCL, London, United Kingdom

## Abstract

**Objective:** To test the hypotheses that blood concentrations of neurofilament light chain protein (NfL) and glial fibrillary acidic protein (GFAp) can serve as biomarkers for disease severity in COVID-19 patients.

**Methods:** Forty-seven inpatients with confirmed COVID-19 had blood samples drawn on admission for assessing serum biomarkers of CNS injury by Single molecule array (Simoa). Concentrations of NfL and GFAp were analyzed in relation to symptoms, clinical signs, inflammatory biomarkers and clinical outcomes. We used multivariate linear models to test for differences in biomarker concentrations in the subgroups, accounting for confounding effects.

**Results:** In total, 21 % (n = 10) of the patients were admitted to an intensive care unit, whereas the overall mortality rate was 13 % (n = 6). Non-survivors had higher serum concentrations of NfL than patients who were discharged alive both in adjusted analyses (p = 2.6 × 10^−7^) and unadjusted analyses (p = 0.001). Serum concentrations of GFAp were significantly higher in non-survivors than survivors in adjusted analyses (p = 0.02). The NfL concentrations in non-survivors increased over repeated measurements, whereas the concentrations in survivors were stable. Significantly higher concentrations of NfL were found in patients reporting fatigue, while reduced concentrations were found in patients experiencing cough, myalgia and joint pain.

**Conclusion:** Increased concentrations of NfL and GFAp in COVID-19 patients on admission may indicate increased mortality risk. Measurement of blood biomarkers for nervous system injury can be useful to detect and monitor CNS injury in COVID-19.

## Introduction

Emerging evidence suggest that respiratory syndrome coronavirus 2 (SARS-CoV-2) infection may affect the nervous system.^1, 2^ Increasing numbers of patients with COVID-19 are reported to have neurological, neuropsychological and neuropsychiatric symptoms and manifestations.^2–5^ Possible mechanisms for nervous system affection in COVID-19 have been suggested, such as direct infection of the nervous system and inflammatory and autoimmune mechanisms.^6–13^ However, the pathobiology is incompletely understood.^7, 14, 15^

Early identification of central nervous system (CNS) manifestations may guide treatment algorithms and thereby improve clinical outcome. Meticulous neurological monitoring is important to assess the frequency and degree of nervous system affections in COVID-19 patients. Blood-based biomarkers for CNS injury, like Neurofilament light chain protein (NfL) and Glial fibrillary acidic protein (GFAp) may be valuable tools for detection and monitoring manifestation during the acute phase. GFAp is an intermediate filament highly expressed in astrocytes, and is increasingly used as a serum biomarker of astrocytic activation/injury.^16^ NfL is a subunit of neurofilaments, which are cylindrical proteins exclusively located in the neuronal axons, that can be measured in blood as a marker of neuronal injury.^17, 18^ In a recent study, neurochemical evidence of neuronal injury and glial activation in patients with moderate and severe COVID-19 infection was demonstrated by assessment of NfL and GFAp.^19, 20^ However, more studies are required to evaluate the usefulness of these biomarkers in COVID-19 patients.

The aim of this study was to explore the association between disease severity in COVID-19 patients and blood concentrations of NfL and GFAp.

## Methods

### Study population

This study includes 47 adult patients (≥18 years old) with COVID-19, as assessed by a positive SARS-CoV-2 PCR test targeting the E-gene on oro- and nasopharyngeal specimens. The patients were consecutively recruited from Oslo University Hospital (n = 26) and Drammen Hospital, Vestre Viken Hospital Trust (n = 21) between March 6 and May 22, 2020 to a clinical cohort study (Norwegian SARS-CoV-2 study; http://ClinicalTrials.gov, number NCT04381819). Clinical information and routine laboratory samples were for most cases collected within 48 hours after hospitalization. Peripheral blood samples were drawn at inclusion, day 2–5 and day 7–10 during hospitalization and repeated later for patients who were hospitalized longer. Using a modified version of the International Severe Acute Respiratory and emerging Infection Consortium (ISARIC)/World Health Organization (WHO) Clinical Characterization Protocol (CCP), clinical and routine data were abstracted from electronic medical records and deposited into an ISARIC (http://isaric.tghn.org) REDCap database (Research Electronic Data Capture, Vanderbilt University, TN, hosted by University of Oxford, UK).

### Sample processing and analyses of biomarkers

Blood samples were collected with 4 mL Vacuette ® (Greiner bio-one International) and processed within one hour by centrifugation at 2000*g* for 10 minutes at room temperature. Serum aliquots were immediately stored at –80^0^C until analysis. Samples were thawed only once during the processing. Measurement of GFAp and NfL in serum samples were performed in the Clinical Neurochemistry Laboratory at the Sahlgrenska University Hospital, Sweden, by board-certified laboratory technicians blind to clinical data. Commercially available single molecule array (Simoa) assays were run on an HD-X Analyzer (Human Neurology 4‐Plex A assay (N4PA advantage kit, 102153), as described by the manufacturer (Quanterix, Billerica, MA). A single batch of reagents was used; intra-assay coefficients of variation were below 10% for all analyses. The results of blood NfL and GFAp concentrations were compared with age-related reference limits established in house from 2.000 healthy control individuals at the Clinical Neurochemistry Laboratory, Sahlgrenska University Hospital, Sweden (unpublished data).

### Statistical analysis

For statistical analyses, the R software with a common set of packages for the purpose was used.^21^ Unique multivariate linear models were used to test for changes in the levels of all biomarkers on admission to address group differences in symptoms, clinical signs and outcomes. Age, gender and creatinine were adjusted for in all linear models separately and unique models of confounding effects according to the resulting performance of the respective linear model were acquired. To correlate between NfL and GFAp concentrations with levels of the other biomarkers, Pearson’s correlations were conducted. The biomarker data were logarithmic transformed to account for the lack of normal distribution. For the biomarkers with low resulting levels (between 0 and 1), a constant of 1 was added to avoid resulting negative log transformed values. All tests were two-sided and P-values < .05 were considered significant.

### Ethical considerations

Informed consents were obtained from all patients or next-of-kin if patients were incapacitated of giving consent. The study was approved by the South-Eastern Norway Regional Health Authority (reference number: 106624).

### Sources of support

This study received funding from Oslo University Hospital and the Research Council of Norway grant no 312780 and has received private donation from Vivaldi Invest A/S owned by Jon Stephenson von Tetzchner.

## Results

### Baseline characteristics

The mean age of the included 47 patients was 60.3 (SD 16.3, range 27–93) years and the male proportion was 72 % (n = 34) (Table 1). On average, the patients had symptoms of COVID-19 infection for nine days (range 0–45) before hospitalization due to COVID-19 disease. The most common neurological symptoms among all patients were headache, ageusia, anosmia and confusion, while none of the patients suffered from stroke (Table 1). Moreover, 26 patients had myalgia (68 % of all reported cases) and 10 patients reported joint pain (26 % of all reported cases). In total, 21 % (n = 10) of the patients were admitted to an intensive care unit (ICU). Six patients (13 %) died from COVID-19 infection during the hospital stay.

**Table 1.** Characteristics of the COVID-19 cohort included in the study.

|  | Baseline |
| --- | --- |
| **(a) Characteristics** | n=47 |
| Female % (n) | 28% (13) |
| Age, mean (SD, range), years | 60.3 (16.3, 27-93) |
| Days from symptom onset until hospitalization (SD, range) | 9.0 (7.7, 0-45) |
| Weight, mean (SD, range), kg | 80.1 (16.7, 54-110) |
| Height, mean (SD, range), cm | 173.8 (11.0, 160-195) |
| BMI, mean (SD, range) | 26.0 (4.6, 18.3-33.8) |
| Present and previous smoking % (n) | 26 (12) |
| Intensive care unit % (n) | 21 (10) |
| **(b) Symptoms and signs on admission** |  |
| History of fever % (n) | 89 (40) |
| Temperature, mean, (SD, range), degrees Celsius | 37.9 (1.0, 35.9-39.8) |
| Cough % (n) | 85 (34) |
| Fatigue % (n) | 19 (8) |
| Anorexia % (n) | 42 (8) |
| **(c) Neurological symptoms** |  |
| Headache % (n) | 37 (14) |
| Ageusia % (n) | 21 (4) |
| Anosmia % (n) | 16 (3) |
| Confusion % (n) | 13 (6) |
| Seizures % (n) | 2 (1) |
| Meningitis / encephalitis % (n) | 5 (1) |
| Neurological disorders % (n) | 4 (2) |
| Stroke % (n) | 0 (0) |
| **(d) Musculoskeletal symptoms** |  |
| Myalgia % (n) | 68 (26) |
| Joint pain % (n) | 26 (10) |
| **(e) Biomarkers on admission** |  |
| Serum GFAP concentrations, mean (SD, range), pg/mL | 286.4 (221, 74-1212) |
| Above cut-off, % (n) | 48 (22) |
| Serum NfL concentrations, mean (SD, range), pg/mL | 33.7 (36.0, 5.8-174.4) |
| Above cut-off, % | 30 (14) |
| CRP, mean (SD, range), mg/L | 97.4 (92.4, 0-400) |
| Ferritin, mean (SD, range), µg/L | 952 (747, 21-3465) |
| White blood cell count, mean (SD, range), x 10<sup>9</sup>/L | 6.5 (3.1, 2.6-18.0) |
| Procalcitonin, mean (SD, range), µg/L | 0.7 (0.1, 0-16.3) |
| CK, mean (SD, range), U/L | 331.9 (733.4, 19-3572) |
| Creatinine, mean (SD, range), µmol/L | 95.8 (51.4, 55-281) |
| Neutrophil granulocyte count, mean (SD, range), x 10<sup>9</sup>/L | 4.8 (27, 1.3-11.3) |

### Serum concentrations of NfL and GFAp in COVID-19 patients

On admission, NfL and GFAp concentrations above reference limits were measured in 30 % (n = 14) and 48 % (n = 22) of the COVID-19 patients, respectively (Table 1). Strong correlations between NfL concentrations and GFAp (p = 2.2 × 10^−7^), procalcitonin-(p = 0.001) and creatinine (p< 0.001) concentrations and neutrophil granulocyte count (p = 0.01) were found. No correlation between NfL concentrations and CRP, creatine kinase, ferritin or white blood cell count was detected (Figure 1). GFAp concentrations were not associated with any other biomarkers than NfL concentrations (Figure 2). When comparing differences in NfL and GFAp concentrations to symptoms, we found that NfL was significantly higher among patients with fatigue (p = 0.02) and reduced in patients with myalgia (p = 8.7 × 10^−4^), cough (p = 3.1 × 10^−3^) and with joint pain (p = 6.6 × 10^−3^) (Table 2). However, no associations between GFAp concentrations and clinical signs were found.

**Figure 1.**
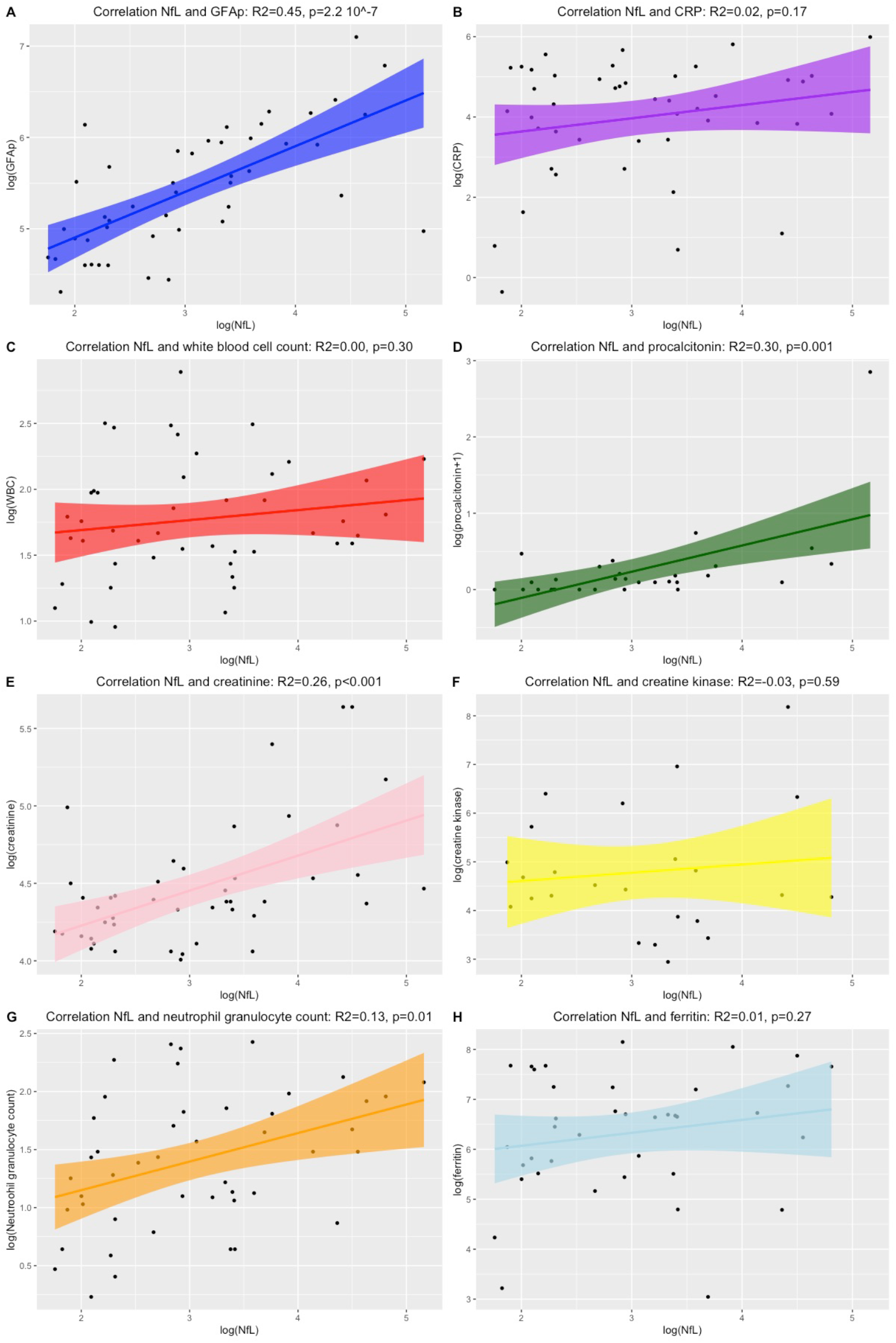
An overview of Pearson’s correlation between NfL concentrations and other biomarkers. Depicted are the correlations between NfL and GFAp concentrations (A), CRP (B), white blood cell count (C), procalcitonin (D), creatinine (E), creatine kinase (F), neutrophil granulocyte count (G) and ferritin (H). Depicted are the logarithmic transformed values.

**Figure 2.**
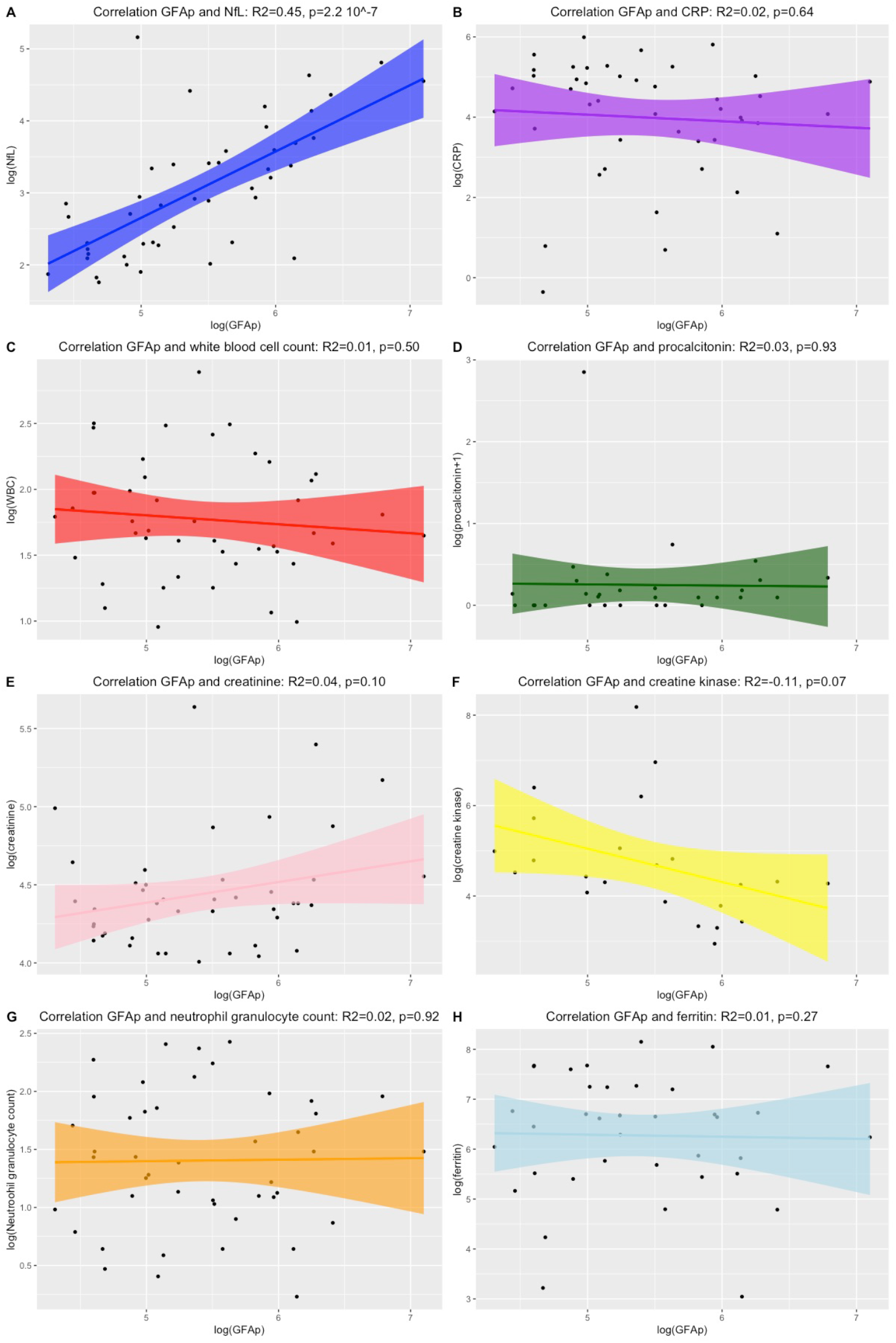
An overview of Pearson’s correlation between GFAp concentrations and other biomarkers. Depicted are the correlations between GFAp concentrations and NfL concentrations(A), CRP (B), white blood cell count (C), procalcitonin (D), creatinine (E), creatine kinase (F), neutrophil granulocyte count (G) and ferritin (H). Depicted are the logarithmic transformed values.

**Table 2.** Differences in NfL concentrations related to symptoms, treatment and outcome.

| Symptom | Linear regression, adjusting for age and creatinine ($R^2=0.26$) | | |
| --- | --- | --- | --- |
| | *t* | $R^2$ | *p* |
| Fever | -0.37 | 0.24 | 0.72 |
| **Cough** | 3.16 | 0.38 | **$3.1 \times 10^{-3}$** |
| **Fatigue** | 2.50 | 0.34 | **0.02** |
| Anorexia | -1.24 | 0.46 | 0.23 |
| Headache | 1.73 | 0.29 | 0.09 |
| Ageusia | -1.54 | 0.48 | 0.14 |
| Anosmia | 0.06 | 0.41 | 0.95 |
| Confusion | -0.95 | 0.26 | 0.35 |
| **Myalgia** | 3.59 | 0.42 | **$8.7 \times 10^{-4}$** |
| **Joint pain** | 2.86 | 0.37 | **$6.6 \times 10^{-3}$** |
| Present and/or previous smoking | -0.34 | 0.49 | 0.73 |
| Intensive care unit | -1.93 | 0.30 | 0.06 |
| Ventilatory support | 1.40 | 0.28 | 0.17 |
| **Outcome - Died** | -6.13 | 0.60 | **$2.6 \times 10^{-7}$** |

### Serum concentrations of NfL and GFAp in relation to outcomes

Concentrations of NfL were significantly higher in non-survivors (n = 6) compared to survivors (p = 2.6 × 10^−7^) when adjusting for age and creatinine levels on admission and in unadjusted analyses (p = 0.001) (Figure 3). Additionally, concentrations of GFAp were significantly higher in non-survivors than survivors when adjusting for age (p = 0.02) (Table 3).

**Table 3.**
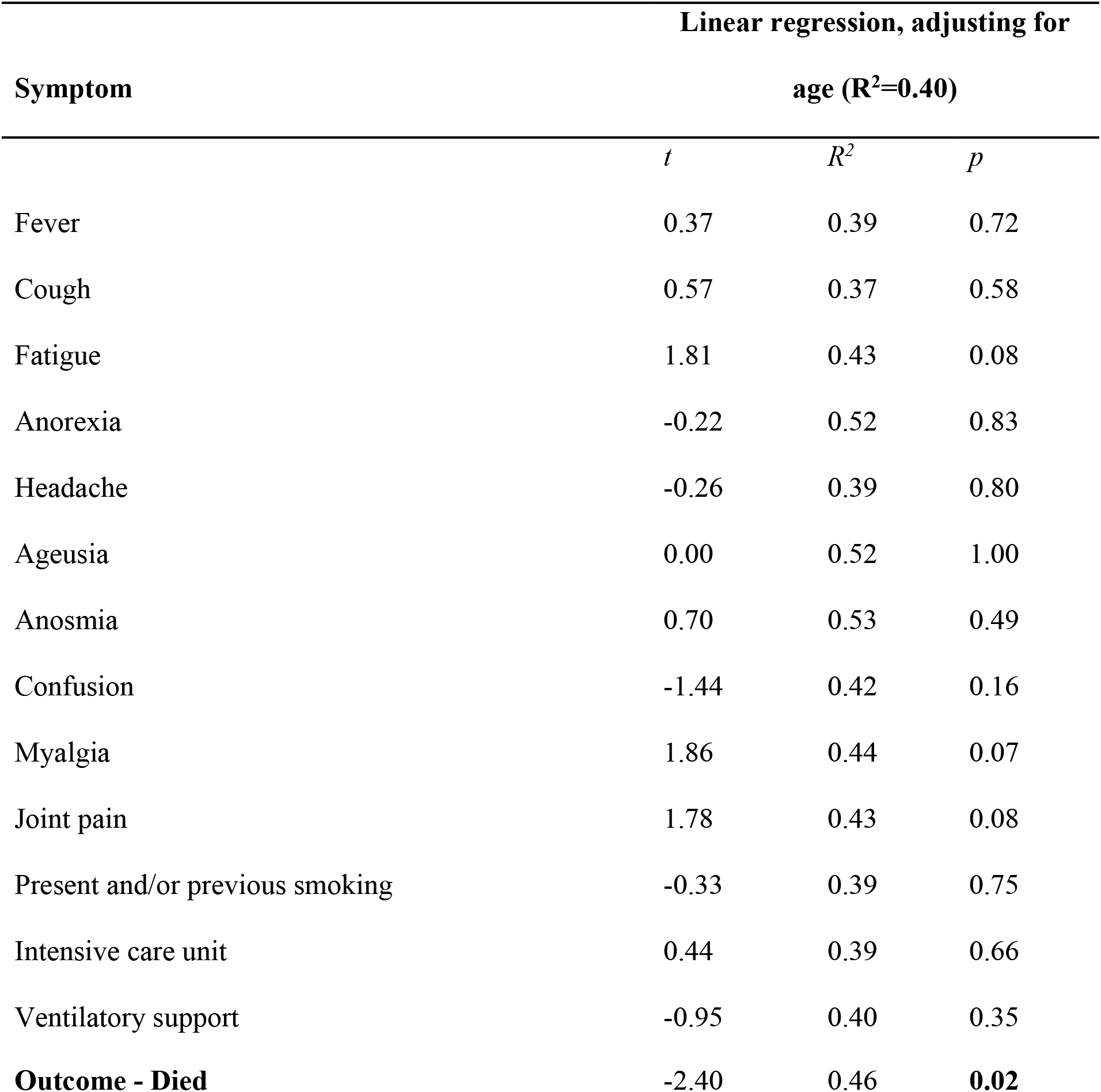
Differences in GFAp concentrations related to symptoms, treatment and outcome.

| Symptom | Linear regression, adjusting for age ($R^2=0.40$) | | |
| --- | --- | --- | --- |
|  | *t* | *R*<sup>2</sup> | *p* |
| Fever | 0.37 | 0.39 | 0.72 |
| Cough | 0.57 | 0.37 | 0.58 |
| Fatigue | 1.81 | 0.43 | 0.08 |
| Anorexia | -0.22 | 0.52 | 0.83 |
| Headache | -0.26 | 0.39 | 0.80 |
| Ageusia | 0.00 | 0.52 | 1.00 |
| Anosmia | 0.70 | 0.53 | 0.49 |
| Confusion | -1.44 | 0.42 | 0.16 |
| Myalgia | 1.86 | 0.44 | 0.07 |
| Joint pain | 1.78 | 0.43 | 0.08 |
| Present and/or previous smoking | -0.33 | 0.39 | 0.75 |
| Intensive care unit | 0.44 | 0.39 | 0.66 |
| Ventilatory support | -0.95 | 0.40 | 0.35 |
| **Outcome - Died** | **-2.40** | **0.46** | **0.02** |

Significant differences among non-survivors compared to survivors were also observed in the adjusted linear models for the level of CRP (p = 0.02), creatine kinase (p = 0.02) and procalcitonin (p = 0.003) on admission, but was not observed for the other biomarkers (white cell blood count, creatinine concentration or neutrophil granulocyte count) (Figure 2 and 3). The longitudinal measurements of NfL concentrations in patients available for repeated measurements, showed increased serum concentrations of NfL at hospital admittance and a tendency of further increased concentrations during hospitalization in patients who died of COVID-19 (Figure 4).

**Figure 3.**
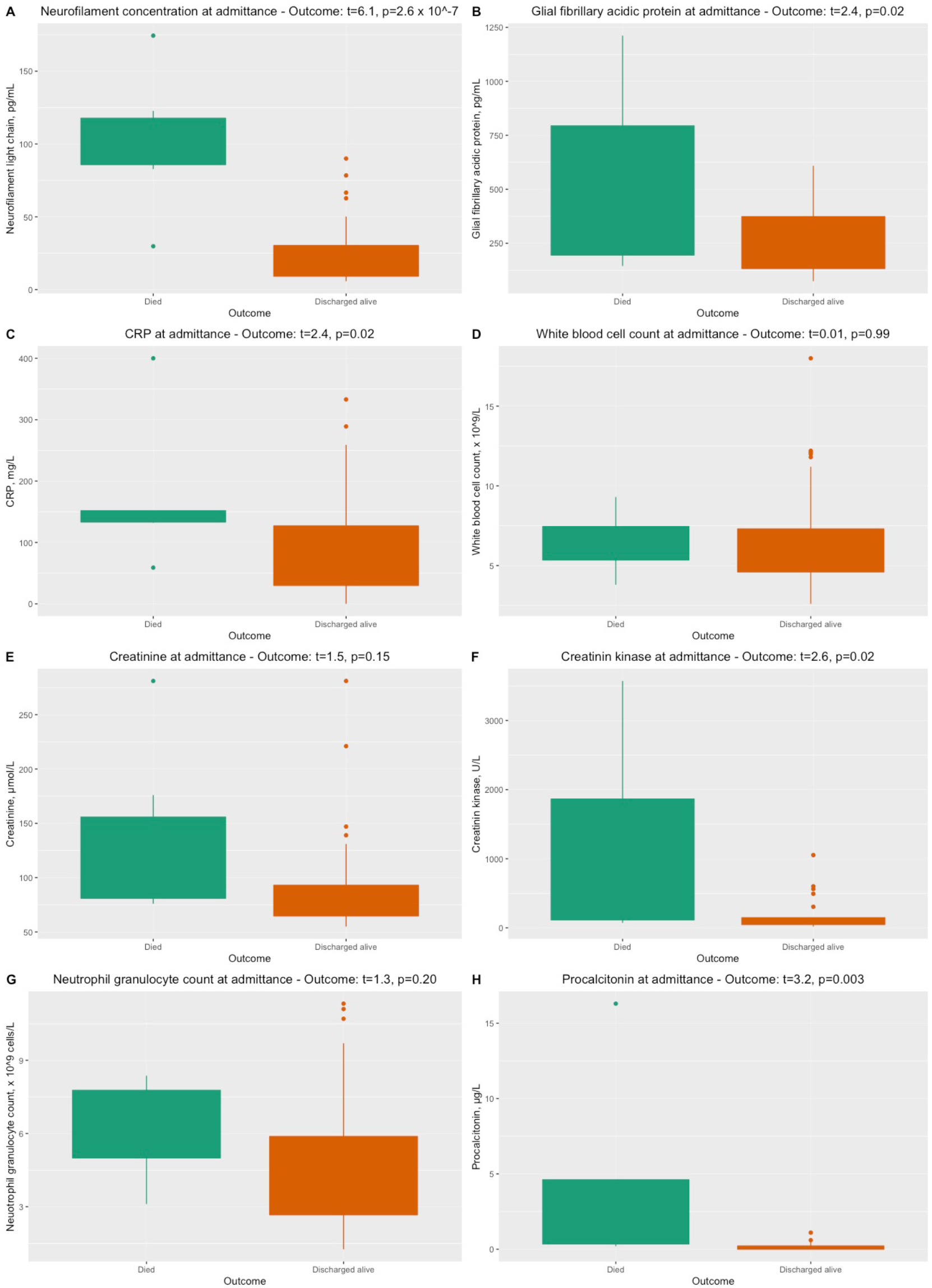
Levels of biomarkers among patients who died and who survived COVID-19 in this study. Statistical analyses performed with unique linear models adjusting for confounding effects. A: NfL concentrations, B: GFAp concentrations, C: CRP, D: White cell blood count, E: creatinine, F: creatine kinase, G: neutrophil granulocyte count and H: procalcitonin.

**Figure 4.**
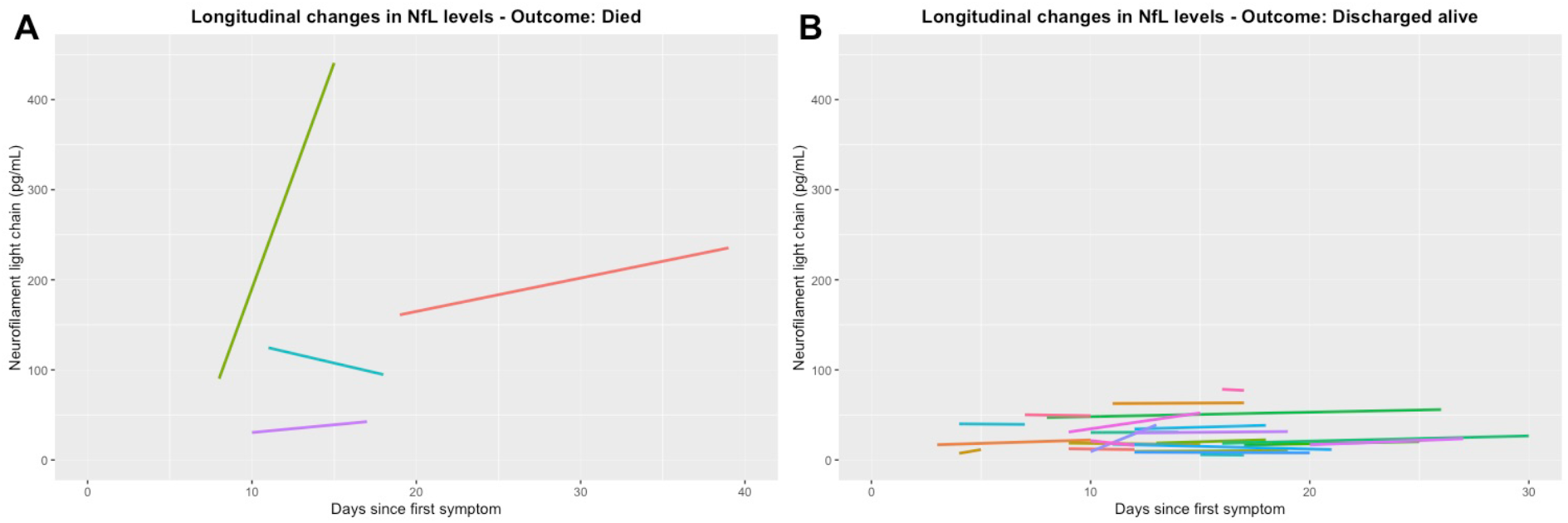
Longitudinal assessment of NfL concentrations among patients who died and who survived COVID-19 in this study. A: Four subjects with longitudinal data who died. B: An overview of the subjects who were discharged alive after hospitalization. Only subjects with longitudinal data are depicted.

The patients with the highest concentrations of NfL (> 120, max 464 pg/mL) were admitted 4 to 7 days after symptom onset and died during their hospital stay (median 14, range 11–42 days after admission). They had respiratory symptoms and malaise as initial symptoms. In addition, they all reported neurological symptoms (headache, dizziness) and a very severe disease course resulting in death during hospitalization. The concentrations of NfL generally increased during the disease course in these subjects.

## Discussion

This pilot study confirms the frequent observations of neurological symptoms in COVID-19 disease. The biomarker results indicate that increased concentrations of serum NfL in patients with COVID-19 may be a predictor of a severe disease course and increased mortality. Concentrations of serum GFAp were also significantly associated with mortality, although not with any CNS symptoms or clinical signs. Increased NfL and GFAp concentration in patients with COVID-19 can be presumed to reflect affection of the nervous system. Although both the peripheral and central nervous system contain NfL, the correlation between cerebrospinal fluid and blood is so high that the majority of the blood NfL must come from the CNS.^22, 23^ Furthermore, GFAp is considered to be fairly specific to CNS.^24^ NfL concentrations in serum at admittance were also associated with reported symptoms of fatigue, which is a general and unspecific symptom.

Of other biomarkers available in this study, increased levels of procalcitonin were apparently associated with increased concentrations of NfL. However, this result is influenced by a few patients with very high measurements. Thus, the implications of these findings are not clear. Moreover, increased levels of creatinine and neutrophil granulocyte counts were associated with higher concentrations of NfL, but the increased concentrations of NfL were not influenced by the creatinine levels in our analyses. Interestingly, NfL concentrations were not correlated with CRP or ferritin, often found to be associated with hyperinflammation in COVID-19 patients, suggesting that the raised NfL concentrations merely reflect enhanced inflammation.

The identification of biomarkers in blood to assess nervous system manifestation will be important in order to monitor the severity of the disease and optimize treatment in COVID-19 patients. Measurements of NfL and GFAp in blood can be clinically useful to assess neurological affection in COVID-19, since this can easily be managed despite medical isolation procedures. Although NfL has been shown to be useful as diagnostic, prognostic and monitoring biomarker in a wide range of other neurological conditions,^23, 25–27^ more studies are needed to assess the applicability of NfL in COVID-19.

One could claim that the high concentrations of NfL reflect medications used in ICU. However, a recent study of NfL and other blood biomarkers in patients undergoing inhalation general anesthesia showed a decrease in NfL concentrations after five hours compared to baseline. This may indicate that increase in NfL concentration in COVID-19 patients treated in ICU might be even larger in magnitude, but are masked by anesthesia-induced decreases.^28^

NfL was not increased in anosmia in this study, although the anosmia in COVID-19 is considered to be related to injury of the olfactory nerve.^11, 29^ Likewise, headache was not associated with increased concentrations of NfL or GFAp in our study. Headache occurring in temporal association with systemic viral infections such as COVID-19, in the absence of meningitis or encephalitis, is well-known clinically.^15, 30^ However, the patients with highest NfL values did all present with headache. They were all intubated shortly after admission, therefore further neurological examination and evaluation was limited. In contrast to other COVID-19 studies, none of the included patients in this study underwent a stroke during hospitalization.^4, 5^

In this pilot study there are several limitations. First, the number of patients with full data sets available was modest. Secondly, detailed and systematic neurological, neurophysiological and neuroradiological investigations were not possible to perform, since our patients were treated under medical isolation procedures at different units and several patients needed ventilatory support in ICUs. Thus, possible association between GFAp and NfL and specific CNS manifestations may have been undetected in this study. In order to study further the questions raised by our observations, we plan a follow-up study of COVID-19 patients up to a year after diagnosis including a systematic neurological assessment.

In conclusion, elevated concentrations of NfL and GFAp in COVID-19 patients seem to be potential prognostic markers in COVID-19. Further studies will be essential in order to elucidate the pathogenesis and the clinical importance of the COVID-19 disease affecting the nervous system and how this can be measured and treated. Prospective neurological and cognitive assessment of individuals with COVID-19 will also be crucial to understand the natural history of COVID-19 in the CNS and monitor for any long-term neurological sequelae.^31^

## Data Availability

The data that support the findings of this study are available on request from the corresponding author, AHA. The data are not publicly available due to ethical restrictions.

## Search terms

SARS-CoV-2, COVID-19, CNS, neurofilament light, glial fibrillary acidic protein, mortality

## Abbreviations used in this paper

CNS: central nervous system;
SARS-CoV-2: severe acute respiratory syndrome coronavirus 2;
ACE-2: angiotensin-converting enzyme 2;
NfL: neurofilament light protein;
GFAp: glial fibrillary acidic protein

## Statistical Analysis

Einar A. Høgestøl and Trine H. Popperud.

## Acknowledgments

We thank all the patients participating in our studies.

## Appendix 1 Authors

| Name | Location | Contribution |
| --- | --- | --- |
| Anne Hege Aamodt,<br>M.D., Ph.D. | Oslo University Hospital, 0424<br>Oslo, Norway | Drafting the manuscript, study<br>concept, design, analyses,<br>interpretation |
| Einar A. Høgestøl,<br>M.D. | University of Oslo and Oslo<br>University Hospital, 0424 Oslo,<br>Norway, Norway | Study design, analyses and<br>interpretation, drafting and<br>approval the manuscript. |
| Trine Haug Popperud, M.D., Ph.D. | Oslo University Hospital, 0424 Oslo, Norway | Design of the work, data analysis and interpretation, drafting the article, final approval of the version to be published. |
| Jan Cato Holter, M.D., Ph.D. | University of Oslo and Oslo University Hospital, 0424 Oslo, Norway, Norway | Design of the work, data analysis and interpretation, drafting the article, final approval of the version to be published. |
| Anne Ma Dyrhol-Riise, M.D., Ph.D.; | University of Oslo and Oslo University Hospital, 0424 Oslo, Norway, Norway | Patient inclusion, interpretation, revision of manuscript, final approval of the version to be published. |
| Kristian Tonby, M.D, Ph.D | University of Oslo and Oslo University Hospital, 0424 Oslo, Norway, Norway | Patient inclusion, revision of manuscript, final approval of the version to be published. |
| Birgitte Stiksrud, MD, PhD | Oslo University Hospital, 0424 Oslo, Norway | Patient inclusion, revision of manuscript, final approval of the version to be published. |
| Else Quist-Paulsen,<br>M.D., Ph.D | Oslo University Hospital, 0424<br>Oslo, Norway | Biochemical measurements,<br>interpretation, revision and final<br>approval of the version to be<br>published. |
| Tone Berge, M.Sc.,<br>Ph.D. | Oslo Metropolitan University,<br>Oslo Norway and Oslo University<br>Hospital, 0424 Oslo, Norway | Study design, data<br>interpretation, revision and final<br>approval of the version to be<br>published. |
| Andreas Barratt-Due,<br>M.D., Ph.D. | Oslo University Hospital, 0424<br>Oslo, Norway | Study concept, data<br>interpretation, revision and<br>final approval of the version to<br>be published. |
| Pål Aukrust, M.D.,<br>Ph.D. | University of Oslo and Oslo<br>University Hospital, 0424 Oslo,<br>Norway, Norway | study concept, data<br>interpretation, revision and<br>final approval of the version to<br>be published. |
| Lars Heggelund, M.D.,<br>Ph.D. | Vestre Viken Hospital Trust,<br>Drammen Hospital,<br>Dronninggata 28, 3004<br>Drammen, Norway | Patient inclusion, revision and<br>final approval of the version to<br>be published. |
| Kaj Blennow, MD,<br>PhD | Clinical Neurochemistry<br>Laboratory, Sahlgrenska<br>University Hospital and<br>Department of Psychiatry and<br>Neurochemistry, the Sahlgrenska<br>Academy at the University of<br>Gothenburg, S-431 80 Mölndal,<br>Sweden | Biochemical measurements,<br>data interpretation, revision and<br>final approval of the version to<br>be published. |
| Henrik Zetterberg,<br>MD, PhD | Department of Psychiatry and<br>Neurochemistry, the Sahlgrenska<br>Academy at the University of<br>Gothenburg, S-431 80 Mölndal,<br>Sweden | Biochemical measurements,<br>data interpretation, revision and<br>final approval of the version to<br>be published. |
| Hanne Flinstad Harbo,<br>M.D., Ph.D, M.H.A. | University of Oslo and Oslo<br>University Hospital, 0424 Oslo,<br>Norway | Study concept, design,<br>analyses, interpretation,<br>drafting and approval of<br>manuscript. |

## References

1. Zhou Z, Kang H, Li S, Zhao X. Understanding the neurotropic characteristics of SARS-CoV-2: from neurological manifestations of COVID-19 to potential neurotropic mechanisms. J Neurol 2020.

2. Wu Y, Xu X, Chen Z, et al. Nervous system involvement after infection with COVID-19 and other coronaviruses. Brain Behav Immun 2020.

3. Heneka MT, Golenbock D, Latz E, Morgan D, Brown R. Immediate and long-term consequences of COVID-19 infections for the development of neurological disease. Alzheimers Res Ther 2020;12:69.

4. Ferrarese C, Silani V, Priori A, et al. An Italian multicenter retrospective-prospective observational study on neurological manifestations of COVID-19 (NEUROCOVID). Neurol Sci 2020;41:1355–1359.

5. Mao L, Jin H, Wang M, et al. Neurologic Manifestations of Hospitalized Patients With Coronavirus Disease 2019 in Wuhan, China. JAMA Neurol 2020.

6. Ahmed MU, Hanif M, Ali MJ, et al. Neurological Manifestations of COVID-19 (SARS-CoV-2): A Review. Front Neurol 2020;11:518.

7. Baig AM, Khaleeq A, Ali U, Syeda H. Evidence of the COVID-19 Virus Targeting the CNS: Tissue Distribution, Host-Virus Interaction, and Proposed Neurotropic Mechanisms. ACS Chem Neurosci 2020;11:995–998.

8. Poyiadji N, Shahin G, Noujaim D, Stone M, Patel S, Griffith B. COVID-19-associated Acute Hemorrhagic Necrotizing Encephalopathy: CT and MRI Features. Radiology 2020:201187.

9. Coen M, Jeanson G, Culebras Almeida LA, et al. Guillain-Barre syndrome as a complication of SARS-CoV-2 infection. Brain Behav Immun 2020.

10. Zanin L, Saraceno G, Panciani PP, et al. SARS-CoV-2 can induce brain and spine demyelinating lesions. Acta Neurochir (Wien) 2020.

11. Netland J, Meyerholz DK, Moore S, Cassell M, Perlman S. Severe acute respiratory syndrome coronavirus infection causes neuronal death in the absence of encephalitis in mice transgenic for human ACE2. J Virol 2008;82:7264–7275.

12. Moriguchi T, Harii N, Goto J, et al. A first case of meningitis/encephalitis associated with SARS-Coronavirus-2. Int J Infect Dis 2020;94:55–58.

13. Wang HY, Li XL, Yan ZR, Sun XP, Han J, Zhang BW Potential neurological symptoms of COVID-19. Ther Adv Neurol Disord 2020;13:1756286420917830.

14. Aamodt AH, Flinstad Harbo H, Eldoen G, Barratt-Due A, Aukrust P. How does COVID-19 affect the brain? Tidsskr Nor Laegeforen 2020;140.

15. Belvis R. Headaches During COVID-19: My Clinical Case and Review of the Literature. Headache 2020.

16. McMahon PJ, Panczykowski DM, Yue JK, et al. Measurement of the glial fibrillary acidic protein and its breakdown products GFAP-BDP biomarker for the detection of traumatic brain injury compared to computed tomography and magnetic resonance imaging. J Neurotrauma 2015;32:527–533.

17. Zetterberg H, Blennow K. Fluid biomarkers for mild traumatic brain injury and related conditions. Nat Rev Neurol 2016;12:563–574.

18. Giovannoni G. Peripheral blood neurofilament light chain levels: the neurologist’s C-reactive protein? Brain 2018;141:2235–2237.

19. Kanberg N, Ashton NJ, Andersson LM, et al. Neurochemical evidence of astrocytic and neuronal injury commonly found in COVID-19. Neurology 2020.

20. Virhammar J, Kumlien E, Fallmar D, et al. Acute necrotizing encephalopathy with SARS-CoV-2 RNA confirmed in cerebrospinal fluid. Neurology 2020.

21. R: A Language and Environment for Statistical Computing [computer program]. Vienna, Austria: R Foundation for Statistical Computing, 2017.

22. Gaetani L, Blennow K, Calabresi P, Di Filippo M, Parnetti L, Zetterberg H. Neurofilament light chain as a biomarker in neurological disorders. J Neurol Neurosurg Psychiatry 2019;90:870–881.

23. Disanto G, Barro C, Benkert P, et al. Serum Neurofilament light: A biomarker of neuronal damage in multiple sclerosis. Ann Neurol 2017;81:857–870.

24. Thelin EP, Zeiler FA, Ercole A, et al. Serial Sampling of Serum Protein Biomarkers for Monitoring Human Traumatic Brain Injury Dynamics: A Systematic Review. Front Neurol 2017;8:300.

25. Gafson AR, Barthelemy NR, Bomont P, et al. Neurofilaments: neurobiological foundations for biomarker applications. Brain 2020.

26. Verde F, Steinacker P, Weishaupt JH, et al. Neurofilament light chain in serum for the diagnosis of amyotrophic lateral sclerosis. J Neurol Neurosurg Psychiatry 2019;90:157–164.

27. Bagnato S, Grimaldi LME, Di Raimondo G, et al. Prolonged Cerebrospinal Fluid Neurofilament Light Chain Increase in Patients with Post-Traumatic Disorders of Consciousness. J Neurotrauma 2017;34:2475–2479.

28. Deiner S, Baxter MG, Mincer JS, et al. Human plasma biomarker responses to inhalational general anaesthesia without surgery. Br J Anaesth 2020.

29. Izquierdo-Dominguez A, Rojas-Lechuga MJ, Mullol J, Alobid I. Olfactory dysfunction in the COVID-19 outbreak. J Investig Allergol Clin Immunol 2020:0.

30. Bolay H, Gul A, Baykan B. COVID-19 is a Real Headache! Headache 2020.

31. Frontera J, Mainali S, Fink EL, et al. Global Consortium Study of Neurological Dysfunction in COVID-19 (GCS-NeuroCOVID): Study Design and Rationale. Neurocrit Care 2020.

